# Pattern of Cutaneous Neoplasms and Associated Factors at a Tertiary Teaching Hospital Pathology Center in Ethiopia: An Eight-Year Histopathological Review

**DOI:** 10.1101/2023.08.26.23294667

**Authors:** Fuad Temam, Samia Metena Yahya, Bereket Berhane, Frehiwot Daba, Amanuel Yeneneh Teka, Indris Ahmed Yesuf, Tigist Workneh Leulseged

## Abstract

**Background:** Cancer is the leading cause of death globally and is on the rise in Africa. Cutaneous neoplasms are becoming increasingly common worldwide. Understanding the patterns of this disease is essential for developing data-driven preventive, screening, and treatment services. However, there are limited studies in Ethiopia so far. Therefore, the study aimed to assess the pattern and associated factors of cutaneous neoplasm among patients with histopathologically confirmed biopsy results at a tertiary teaching hospital in Ethiopia from March, 2014 to October, 2022.

**Methods:** A retrospective record review study was conducted among 1006 patients with histopathologically confirmed cutaneous neoplasms from the biopsies that were assessed at St. Paul’s Hospital Millennium Medical College. Data was summarized using frequencies (percentages), median (interquartile range), and graphs. To identify significant factors associated with malignant cutaneous neoplasm, a multivariable binary logistic regression model was fitted, where Adjusted Odds ratio (AOR), 95% CIs for AOR, and p-values were used for interpretation of results.

**Result:** From the 1006 cases, 265 (26.3%, 95%CI=23.5%-29.3%) were malignant, of which sarcoma (26.0%) and squamous cell carcinoma (25.7%) were the most frequent and found to be prevalent in younger (19-29 years) and older (≥60 years) patients, respectively. The trunk was the commonest site (54.2%) for all the malignancies, especially sarcoma (80.4%). Age was found to be a significant exposure that is associated with the development of malignant cutaneous neoplasm for those ≥30 years as compared with those ≤18 years, with the odds increasing with age (AOR=2.66, 95% CI=1.10,6.45 for 30-39 years, AOR=4.98, 95% CI= 2.01,12.36 for 40-49 years, AOR=5.33, 95% CI=2.15,13.22 for 50-59 years and AOR= 6.62, 95% CI=2.79,15.66 for ≥60).

**Conclusion:** The prevalence of malignant cutaneous neoplasm is higher than previously reported in the country and the malignancy pattern and distribution are different from what is known so far. This could signal a shift in disease epidemiology, and the findings should be factored into clinical decision making and program design for disease prevention, screening, and treatment. It also calls for further prospective research to learn more about the condition in the context of additional relevant personal and clinical characteristics.

## BACKGROUND

Neoplasms are abnormal tissue growths that continue to grow in the absence of external stimuli. They can be benign, in which the growth does not invade nearby tissues or other parts of the body, or malignant (also called cancers), in which the growth can invade nearby tissues and has the potential to metastasize to other parts of the body via the blood and lymph systems [1-3].

According to WHO, cancer has long been the top cause of mortality worldwide, accounting for millions of deaths before the age of 70 in most countries, with an estimated 19.3 million new cancer cases and over 10.0 million cancer deaths in 2020 [4-8]. Cancer incidence in Africa has also increased from 715,000 in 2008 to 1.1 million in 2020, and associated mortality has increased from 542,000 in 2008 to 711,000 in 2020 [9,10]. In 2019, the national burden of cancer in Ethiopia was estimated to be 53,560 new cases, 39,480 deaths, and 1.42 million disability-adjusted life years (DALYs) lost due to cancer [11]. These estimates are supported by studies undertaken at different tertiary level institutions in the country [12-17].

Cutaneous neoplasms, one type of neoplasm, are abnormal growths on the skin that can be either benign or malignant. The most common types of cutaneous neoplasms are basal cell carcinoma (BCC), squamous cell carcinoma (SCC), and melanoma. Cutaneous neoplasms are among the most common forms of cancer, and the global incidence has been increasing at an alarming rate [1,18,19]. Understanding the pattern of this disease is essential for developing data-driven preventive, screening, and treatment services. However, the area is under-researched in Africa, despite the fact that many studies have been conducted around the world [10, 20-22]. In Ethiopia, there have been a few studies, but they have all focused on malignant cutaneous neoplasms. These studies have found that sarcomas account for 9.7% to 15% of all neoplasms, and SCC accounts for 8% of all cutaneous biopsy specimens [12, 13, 16, 17].

This study aims to fill this knowledge gap by examining the pattern of both benign and malignant cutaneous neoplasms in one of the largest referral tertiary hospitals in Ethiopia, which receives a large number of patients from all over the country, and has a well-established pathology center. Accordingly, the study aimed to assess the pattern and associated factors of cutaneous neoplasms among patients with histopathologically confirmed biopsy results at a tertiary teaching hospital in Ethiopia between March 2014 and October 2022.

## METHODOLOGY

### Study Setting and Period

The study was conducted from December 2022 to February 2023 at St Paul’s Hospital Millennium Medical College (SPHMMC), a tertiary teaching hospital under the Federal Ministry of Health in Addis Ababa, Ethiopia. The hospital provides both inpatient and outpatient service under different specialty and subspecialty fields to more than 5 million population under its catchment. Among its well-established wings is the Pathology center, which started service in 2014. The center is one of the few Pathology centers in the country. It archives specimens from many different other institutions, and provides FNAC service for more than 7,000 patients and biopsy service for an average of 6000 patients annually.

### Study Design and Population

A retrospective chart review study was conducted among patients who had histopathological biopsy specimens examined at the pathology center between March 2014 and October 2022.

The source population were all patients for whom histopathological analysis of dermatology and soft tissue biopsy specimens were made at the pathology center of the hospital during the eight-year observation period. From whom, all patients with confirmed cutaneous neoplasm were the study population. From the list of the study population, patients with complete data on major demographic, clinical and histopathologic data were eligible to be included in the study.

### Sample Size and Sampling

All patients with histopathologically proven cutaneous neoplasm and with complete data on age, sex and anatomic site were included in the study. During the eight-year observation period, a total of 1054 dermatology and soft tissue biopsy specimens of patients were analyzed, from which 1006 eligible patients were included in the study. (**Figure 1**)

**Figure 1:**
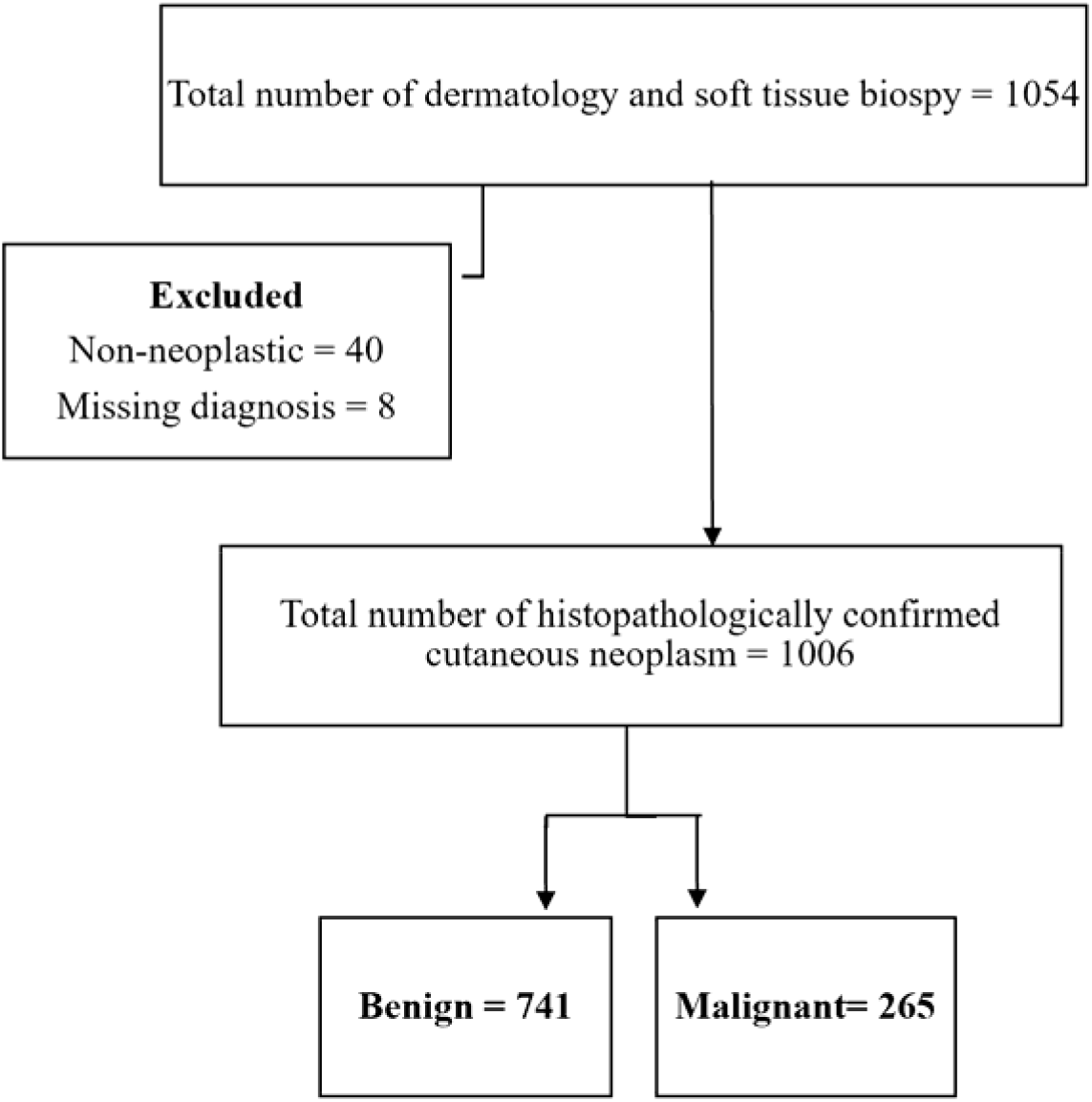
Flow chart showing the selection of study participants included in the study

### Data Collection and Quality Assurance

Data was extracted from the electronic database of the Department of Pathology using a predefined data abstraction tool on the following variables; age, sex, anatomic location of the neoplasm, type of cutaneous neoplasm, and pattern of cutaneous malignancy.

Data extraction was made by two trained General Practitioners. Further data quality was assured by data cleaning through checking for inconsistencies, numerical errors and missing parameters. Furthermore, the patients’ records were referred to verify the collected data whenever discrepancies were observed. All statistical analysis was performed using SPSS version 25.0 software for analysis.

### Statistical Analysis

Data was described with the proper statistical summary measures based on the nature of the variable using frequencies with percentages, graphs and median with interquartile range.

To identify factors associated with the development of malignant cutaneous neoplasm among the study participants, a binary logistic regression model was run. All the exposure variables were included in the multivariable analysis, despite their statistical level at univariate analysis, due to the clinical significance of the variables. The final multivariable model was run at 5% level of significance. To measure the presence and strength of association between the exposures and the type of neoplasm, adjusted odds ratio (AOR), 95% CI for AOR and P-value was used. Variables with p-value ≤ 0.05 were considered as significantly associated with the development of malignant cutaneous neoplasms. The adequacy of the final model was checked using the Hosmer and Lemeshow goodness of fit test and the model fitted the data well with p-value of 0.290.

## RESULTS

### Sociodemographic and clinical characteristics

The median age of the participants was 35.7 years (IQR, 24.9 - 54.0), of which 121 (12.0%) were pediatric cases, and 518 (51.5%) were females. Two hundred sixty-five (26.3%, 95% CI= 23.5% -29.3%) of the cases were malignant and the rest 741 (73.7%, 95% CI=70.7% - 76.5%) were benign. (**Table 1**)

**Table 1:** Socio–demographic and clinical characteristics of patients (n=1006)

| Variable | Frequency | Percentage |
| --- | --- | --- |
| **Age category (in years)** |  |  |
| ≤ 18 | 121 | 12.0 |
| 19-29 | 246 | 24.5 |
| 30-39 | 196 | 19.5 |
| 40-49 | 129 | 12.8 |
| 50-59 | 119 | 11.8 |
| ≥ 60 | 195 | 19.4 |
| **Sex** |  |  |
| Male | 488 | 48.5 |
| Female | 518 | 51.5 |
| **Type of neoplasm** |  |  |
| Benign | 741 | 73.7 |
| Malignant | 265 | 26.3 |

From these 265 malignant cases, 212 (80%) is constituted by five malignancies; sarcoma (26.0%) and squamous cell carcinoma (25.7%) being the most frequently diagnosed malignancies followed by melanoma (12.1%), basal cell carcinoma (9.4%), and lymphoma (6.8%). Among the 69 sarcoma cases nearly half (31/69) was dermatofibrosarcoma protuberans. Pleomorphic sarcoma and leiomyosarcoma accounted for 5/69 cases each, Kaposi sarcoma was diagnosed in 4/69, and low grade fibromyxoid sarcoma was diagnosed in 2/69. The remaining 22/69 sarcomas were of different types. From the 1006 cases, data on anatomic site of neoplasm was accessed only for 643 cases. Of these 643 cases, only 8 (1.2%) had multiple site lesions, the rest 635 (98.8%) had lesions on only one anatomic site. Accordingly, the commonest site was trunk constituting more than half (54.2%) of the cases followed by head, neck and face (22.0%), lower limb (18.4%) and upper limb (5.4%). (**Figure 2**)

**Figure 2:**
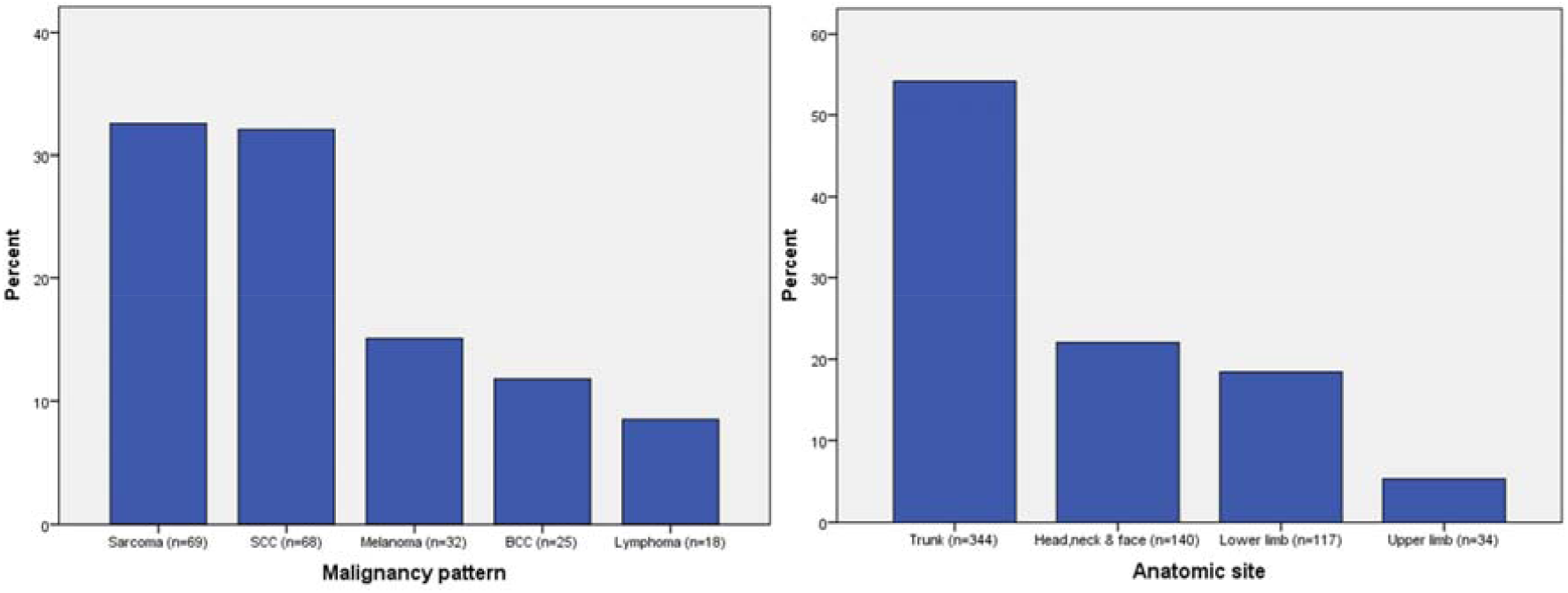
Malignancy pattern and anatomic site of the lesions among patients.

### Distribution of common cutaneous malignancies based on age, sex and anatomic site

The distribution of the commonly identified neoplasms showed that Sarcoma was more frequently diagnosed in those younger than 60 years with the largest proportion lying between 19 and 29 years of age (30.4%). SCC, melanoma and BCC are diagnosed in older age groups notably among those older than 60 years (64.0%, 37.5% and 41.2%, respectively). On the other hand, diagnosis of lymphoma seems to have a bi-modal pick where most diagnosed cases lie in the age range of 19 to 39 (55.5%) and older than 60 years (27.8%). The distribution of these malignancies based on sex doesn’t seem to have a big difference except for SCC where almost two-third (61.8%) of cases were diagnosed in females.

Regarding anatomic sites, the majority (80.4%) of sarcoma cases were located on the trunk. SCC was comparably distributed on all sites except on the upper limb (3.9%). For melanoma, almost all cases were located on the trunk (43.5%) and lower limb (52.2%) and none on the upper limb. The most frequent site for BCC is the head, neck and face region (84.6%) and the rest 2 (15.4%) were located on the Trunk. All cases of lymphoma were on the trunk (40.0%) and limbs (40.0% on upper limb and 20.0% on lower limb). **(Table 2)**

**Table 2:** Distribution of the common cutaneous malignancies based on age, sex and anatomic site.

| Variable | Type of cutaneous malignancy |  |  |  |  |
| --- | --- | --- | --- | --- | --- |
|  | Sarcoma<br>(n=69) | SCC<br>(n=68) | Melanoma<br>(n=32) | BCC<br>(n=25) | Lymphoma<br>(n=18) |
| **Age category**<br>**(n= 212)** |  |  |  |  |  |
| ≤ 18 | 9 (13.0%) | 0 | 1 (3.1%) | 0 | 1 (5.6%) |
| 19-29 | 21 (30.4%) | 2 (2.9%) | 2 (6.3%) | 0 | 4 (22.2%) |
| 30-39 | 9 (13.0%) | 11 (16.2%) | 7 (21.9%) | 1 (4.0%) | 6 (33.3%) |
| 40-49 | 14 (20.3%) | 12 (17.6%) | 6 (18.8%) | 5 (20.0%) | 2 (11.1%) |
| 50-59 | 10 (14.5%) | 15 (22.1%) | 4 (12.5%) | 3 (12.0%) | 0 |
| ≥ 60 | 6 (8.7%) | 28 (41.2%) | 12 (37.5%) | 16 (64.0%) | 5 (27.8%) |
| **Sex (n=212)** |  |  |  |  |  |
| Female | 29 (42.0%) | 42 (61.8%) | 18 (56.3%) | 11 (44.0%) | 8 (44.4%) |
| Male | 40 (58.0%) | 26 (38.2%) | 14 (43.8%) | 14 (56.0%) | 10 (55.6%) |
| **Anatomic site (n=143)** |  |  |  |  |  |
| Head, neck and face | 5 (9.8%) | 20 (39.2%) | 1 (4.3%) | 11 (84.6%) | 0 |
| Trunk | 41 (80.4%) | 17 (33.3%) | 10 (43.5%) | 2 (15.4%) | 2 (40.0%) |
| Upper limb | 1 (2.0%) | 2 (3.9%) | 0 | 0 | 2 (40.0%) |
| Lower limb | 4 (7.8%) | 12 (23.5%) | 12 (52.2%) | 0 | 1 (20.0%) |

### Factors associated with type of cutaneous neoplasm (benign Vs malignant)

To identify factors that affect the type of cutaneous neoplasm, a binary logistic regression model was applied. Multivariable analysis was run at 5% level of significance to identify if the exposures are significantly associated with the diagnosis of malignant cutaneous neoplasm.

Accordingly, age group was found to be the only exposure that is significantly associated with the outcome. After adjusting for sex and anatomic site, the odds of developing malignant cutaneous neoplasm showed a significant increase for those 30 years and older as compared with those 18 years and younger. The increase was higher with increase in age with almost a triple increase among those 60 years and older (AOR= 6.62, 95% CI= 2.79, 15.66, p<0.0001) as compared to those 30-39 years old (AOR= 2.66, 95% CI= 1.10, 6.45, p=0.03). On the other hand, patients 19-29 years old didn’t show a statistically significant increased odds of developing malignant cutaneous neoplasm as compared with those 18 years and younger. (**Table 3**)

**Table 3:** Factors associated with development of malignant cutaneous neoplasm (n=635)

| Variable | COR (95% CI) | AOR (95% CI) | p-value |
| --- | --- | --- | --- |
| **Age category** |  |  |  |
| ≤ 18 | 1 | 1 | <0.0001* |
| 19-29 | 1.93 (0.79, 4.69) | 1.89 (0.78, 4.61) | 0.158 |
| 30-39 | 2.68 (1.11, 6.46) | 2.66 (1.10, 6.45) | **0.03\*** |
| 40-49 | 4.87 (1.97, 12.04) | 4.98 (2.01, 12.36) | **0.001\*** |
| 50-59 | 5.35 (2.17, 13.23) | 5.33 (2.15, 13.22) | **0.000\*** |
| ≥ 60 | 6.38 (2.71, 15.04) | 6.62 (2.79, 15.66) | **0.000\*** |
| **Sex** |  |  |  |
| Male | 1 | 1 |  |
| Female | 1.19 (0.85, 1.68) | 1.33 (0.93, 1.91) | 0.117 |
| **Anatomic site** |  |  |  |
| Head, neck and face | 1 | 1 | 0.413 |
| Trunk | 0.87 (0.57, 1.32) | 0.86 (0.55, 1.34) | 0.495 |
| Upper limb | 0.45 (0.18, 1.17) | 0.46 (0.18, 1.23) | 0.122 |
| Lower limb | 0.87 (0.51, 1.48) | 0.75 (0.43, 1.30) | 0.301 |

**Note:** COR, Crude Risk ratio; AOR, Adjusted Risk ratio; CI, Confidence interval; \*statistically significant

## DISCUSSION

The study was conducted among 1006 patients with histopathologically confirmed cutaneous neoplasms with the aim of assessing the pattern and associated factors of cutaneous neoplasm. Among the total cases, 265 (26.3%, 95% CI= 23.5% -29.3%) of the cases were malignant and the rest 741 (73.7%, 95% CI=70.7% - 76.5%) were benign. The prevalence of malignant cutaneous neoplasms in our study appears to be close to that of another study conducted in Ethiopia, which reported a frequency of 22.0%. However, it is higher than studies from another regional institution in Ethiopia, where a frequency of 4.06% was reported, and Nigeria, where a prevalence of 12.7% was reported [15,16, 20].

The study revealed that sarcoma (26.0%) and SCC (25.7%) were the most frequently diagnosed malignancies followed by melanoma (12.1%), BCC (9.4%), and lymphoma (6.8%). Among the 69 sarcoma cases, nearly half (31/69) were dermatofibrosarcoma protuberans. This is in contrast to most data from Ethiopia, where SCC is the most frequent form of cutaneous malignancy, accounting for about half of all cases [13, 15-17]. Similarly, a study conducted in Kenya found SCC to be the most frequent cutaneous malignancy, accounting for 45% of all cases [21]. Furthermore, the study showed that the commonest anatomic site was the trunk constituting more than half (54.2%) of the cases followed by head, neck and face (22.0%), lower limb (18.4%) and upper limb (5.4%). This is also in contrast to previous findings where lower limb is reported to be the commonest site of cutaneous neoplasm accounting for nearly half of the neoplasms [13, 21].

The difference in the frequency and pattern of malignant cutaneous neoplasms seen in this study as compared to previous researches could be attributed to the following factors: the nature of the institution where the study was conducted, which receives more serious cases that require better medical attention from all over the country, the potentially increased healthcare-seeking behavior of the population in recent years, and a probable change in disease epidemiology due to continuously changing lifestyle and climate change. In addition, the equal prevalence of sarcomas and SCC in our study could be due to the higher submission rate of soft tissue biopsies as compared to skin biopsies based on routine observation. However, this could not be objectively demonstrated as the tissue samples are not submitted as soft tissue and skin separately.

On the regression analysis, age was found to be a significant exposure that is associated with the development of malignant cutaneous neoplasm for those 30 years and older as compared with those 18 years and younger with the odds increasing with age. This is an expected pattern as it is known that the risk of any cancer increases with age due to cellular damage accumulation, weakened immune system, less healthy stem cells, and changes in metabolism. This finding is also in line with previous studies conducted in comparable settings [13,21].

The findings of this study add a significant contribution to the current literature by providing an updated finding from a large referral hospital with a large number of participants from all over the country over an eight-year period. However, due to a lack of queries in the pathology patient reception chart, data on additional relevant personal and clinical information were not included in the study.

## CONCLUSION

The study’s findings show that the prevalence of malignant cutaneous neoplasm is higher than previously reported in the country. Furthermore, the malignancy pattern and distribution are different from what is known so far. This could signal a shift in disease epidemiology, and the findings should be factored into clinical decision making and program design for disease prevention, screening, and treatment. It also calls for further prospective research to learn more about the condition in the context of additional relevant personal and clinical characteristics.

## Data Availability

All relevant data are available upon reasonable request.

## DECLARATION

### Ethical Considerations

The study was conducted after securing ethical clearance from St. Paul’s Hospital Millennium Medical College institutional review board (SPHMMC-IRB). Since the study used secondary data, waiver of consent was also obtained from the SPHMMC-IRB. Medical record number was used for the data collection and personal identifiers of the patient were not used in the research report. Access to the collected information was limited to the research team and confidentiality was maintained throughout the project.

### Availability of data and materials

All relevant data are available upon reasonable request.

### Authors Contribution

SM, FT, BB, FD and AY conceived the study, contributed in interpretation of the findings and revised the manuscript. TWL contributed to the conception, designed the study, performed statistical analysis, and drafted the initial manuscript. IAY contributed in data management and analysis, and draft of the initial manuscript. IAY extracted the data and contributed in data management. All authors approved the final version of the manuscript.

## Acknowledgement

The authors would like to thank St. Paul’s Hospital Millennium Medical College for facilitating the research work.

## REFERENCES

1. Roth GA, Abate D, Abate KH. Global, regional, and national age-sex-specific mortality for 282 causes of death in 195 countries and territories, 1980–2017: a systematic analysis for the Global Burden of Disease Study 2017. Lancet. (2018) 392:1736–88. doi:10.1016/S0140-6736(18)32203-7

2. Harrison’s Principles of Internal Medicine, 20eJ. Larry Jameson, Anthony S. Fauci, Dennis L. Kasper, Stephen L. Hauser, Dan L. Longo, Joseph Loscalzo

3. WHO cancer facts 2022, https://www.who.int/news-room/fact-sheets/detail/cancer

4. Roth GA, Abate D, Abate KH. Global, regional, and national age-sex-specific mortality for 282 causes of death in 195 countries and territories, 1980–2017: a systematic analysis for the Global Burden of Disease Study 2017. Lancet. (2018) 392:1736–88. doi: 10.1016/S0140-6736(18)32203-7

5. Cancer in sub-Saharan Africa in 2020: a review of current estimates of the national burden, data gaps, and future needs Freddie Bray 1, D Maxwell Parkin 2, African Cancer Registry Network Collaborators, Affiliations PMID:35550275 DOI: 10.1016/S1470-2045(22)00270-4

6. National Burden and Trend of Cancer in Ethiopia, 2010–2019: a systematic analysis for Globalburden of disease study Atalel Fentahun Awedew1*, Zelalem Asefa1 & Woldemariam Beka Belay 10.1038/s41598-022-17128-9

7. World Health Organization, Global Health Estimates 2020: Deaths by Cause, Age, Sex, by Country and by Region, 2000-2019. WHO; 2020. Accessed December 11, 2020.

8. Global cancer statistics 2018: GLOBOCAN estimates of incidence and mortality worldwide for 36 cancers in 185 countries Freddie Bray, Jacques Ferlay, Isabelle Soerjomataram, Rebecca L Siegel, Lindsey A Torre, Ahmedin Jemal CA: a cancer journal for clinicians 68 (6), 394–424, 2018

9. Mapping Cancer in Africa: A Comprehensive and Comparable Characterization of 34 Cancer Types Using Estimates From GLOBOCAN 2020 Rajesh Sharma1, Aashima1, Mehak Nanda1, Claudio Fronterre2, Paul Sewagudde3, Anna E. Ssentongo4,5, Kelsey Yenney6, Nina D. Arhin7, John Oh5, ForsterAmponsah-Manu8 andPaddySsentongo4,9 April 2022 | Volume 10 | Article 83983

10. Cancer in sub-Saharan Africa in 2020: a review of current estimates of the national burden, data gaps, and future needs Freddie Bray 1, D Maxwell Parkin 2, African Cancer Registry Network Collaborators, Affiliations 10.1016/S1470-2045(22)00270-4

11. National Burden and Trend of Cancer in Ethiopia, 2010–2019: a systemic analysis for Global Burden of disease studyAtalel Fentahun Awedew, Zelalem Asefa & Woldemariam Beka Beay scientific reports 2022) 12:12736

12. Superficial Malignant Neoplasms in Southwestern Ethiopia: A Cytopathological Approach Mesele Bezabih, M.D. Diagnostic Cytopathology, Vol 31, No 5 3

13. Pattern of Cancer in Tikur Anbessa Specialized Hospital Oncology Center in Ethiopia from 1998 to 2010 Wondemagegnehu Tigeneh Abera Molla, Ayenalem Abreh and Mathwose Assefa Int J Cancer Res Mol Mech Volume 1.1: 16966/2381-3318.103

14. Patterns of cancer in the University of Gondar Hospital, NorthWest Ethiopia B.T. Deressa Annals of Oncology VOLUME 27, SUPPLEMENT 9, DECEMBER 01, 2016

15. Dermatopathology practice in Ethiopia Devon C. Gimbel, Teklu B. Legesse Arch Pathol Lab Med. June 2013;137:798–804

16. SKIN CANCER IN TIGRAY REGION (ETHIOPIA) A Morrone (1) - F Dassoni (2) - V Padovese (3) - O Latini (4) - A Scarabello (5) - A Cristaudo (6) - H Godefay Tropical dermatology June 2019

17. Mesele Bezabih, Patterns in Skin Cancers in Tikur Anbessa Hospital; Ethiop J Health Sci Vol. 1L No 1 January 2001

18. Arnold M, Singh D, Laversanne M, Vignat J, Vaccarella S, Meheus F, Cust AE, de Vries E, Whiteman DC, Bray F. Global Burden of Cutaneous Melanoma in 2020 and Projections to 2040. JAMA Dermatol. 2022 May 1;158(5):495–503. doi: 10.1001/jamadermatol.2022.0160. PMID:35353115; PMCID: PMC8968696.

19. Aiyuan Guo, Xiajing Liu, Heqing Li, Wenwei Cheng, Yexun Song, “The Global, Regional, National Burden of Cutaneous Squamous Cell Carcinoma (1990–2019) and Predictions to 2035”, European Journal of Cancer Care, vol. 2023, Article ID 5484597, 8 pages, 2023. 10.1155/2023/5484597

20. Dermatological malignancies in Kano, Northern Nigeria: a histopathological review O Ochicha, ST Edino, AS Mohammed, AB Umar, Annals of African Medicine Vol.3(4) 2004: 188–191

21. Peter M. Nthumba, Pedro C. Cavadas, and Luis Landin, Primary Cutaneous Malignancies in Sub-Saharan Africa; Ann Plast Surg 2011;66: 313–320)

22. Wright CY, du Preez DJ, Millar DA, Norval M. The Epidemiology of Skin Cancer and Public Health Strategies for Its Prevention in Southern Africa. Int J Environ Res Public Health. 2020 Feb 6;17(3):1017. doi: 10.3390/ijerph17031017. PMID:32041101; PMCID: PMC7037230.

